# Variant-specific burden of SARS-CoV-2 in Michigan: March 2020 through November 2021

**DOI:** 10.1101/2022.03.16.22272497

**Authors:** Joshua G. Petrie, Marisa C. Eisenberg, Adam S. Lauring, Julie Gilbert, Samantha M. Harrison, Peter M. DeJonge, Emily T. Martin

**Affiliations:** Center for Clinical Epidemiology & Population Health, Marshfield Clinic Research Institute, Marshfield, WI; Department of Epidemiology, University of Michigan School of Public Health, Ann Arbor, MI; Departments of Internal Medicine and Microbiology and Immunology, University of Michigan, Ann Arbor, MI; Wisconsin Department of Health Services

**Keywords:** COVID-19, SARS-CoV-2, Incidence, Seroprevalence, Infection, Hospitalization, Death, Michigan

## Abstract

Accurate estimates of total burden of SARS-CoV-2 are needed to inform policy, planning and response. We sought to quantify SARS-CoV-2 cases, hospitalizations, and deaths by age in Michigan. COVID-19 cases reported to the Michigan Disease Surveillance System were multiplied by age and time-specific adjustment factors to correct for under-detection. Adjustment factors were estimated in a model fit to incidence data and seroprevalence estimates. Age-specific incidence of SARS-CoV-2 hospitalization, death, and vaccination, and variant proportions were estimated from publicly available data. We estimated substantial under-detection of infection that varied by age and time. Accounting for under-detection, we estimate cumulative incidence of infection in Michigan reached 75% by mid-November 2021, and over 87% of Michigan residents were estimated to have had ≥1 vaccination dose and/or previous infection. Comparing pandemic waves, the relative burden among children increased over time. Adults ≥80 years were more likely to be hospitalized or die if infected in fall 2020 than if infected during later waves. Our results highlight the ongoing risk of periods of high SARS-CoV-2 incidence despite widespread prior infection and vaccination. This underscores the need for long-term planning for surveillance, vaccination, and other mitigation measures amidst continued response to the acute pandemic.

---

Response to the COVID-19 pandemic has been challenged by rapidly changing circumstances including the continued emergence of SARS-CoV-2 variants and a developing understanding of the breadth and duration of vaccine-induced immunity. Accurate estimates of total burden of SARS-CoV-2 infection are needed to inform policy and ongoing response efforts, and forecasting scenarios as the pandemic continues to evolve. Beyond considerations of current COVID-19 protection strategies, estimates of the total burden of infection are critical to assessing the population-level impact of the pandemic, and ongoing monitoring of indicators of severity like the proportion of infected individuals who become hospitalized or die in post-pandemic outbreaks. As policy-makers seek to update decisions in an environment of shifting vaccination and infection patterns, a better understanding of overall level of population immunity based on best-available surveillance data is needed.

Accurately estimating the total burden of SARS-CoV-2 infection is difficult, however. Public health surveillance systems are challenged by persistent under-detection of cases, particularly for those infections that do not require medical attention. This is a major factor in tracking SARS-CoV-2 infections, where approximately one third may be completely asymptomatic^1,2^. Levels of under-detection are also expected to vary by age of the infected individual and over time related to factors such as testing availability and testing behaviors^3^. Seroprevalence studies can be helpful for surveillance and estimating the total burden of infection, but these studies have limitations of their own^4,5^. Seroprevalance estimates provide a snapshot of past and recent infection that can be difficult to disentangle and that underestimates the true burden of infection due to effects of waning antibody^6^.

The emergence of multiple variants of SARS-CoV-2 since the beginning of the pandemic underscores the need for accurate estimates of disease burden. The state of Michigan has experienced four waves of SARS-CoV-2 transmission during the COVID-19 pandemic through December 2021. Each of these waves has affected the general population and healthcare systems in different ways suggesting changing patterns of infection and severity by age. Michigan is also one of few states that experienced substantial transmission of both the SARS-CoV-2 Alpha (B.1.1.7 lineage) and Delta (B.1.617.2 lineage) variants^7^. We sought to quantify the burden of SARS-CoV-2 cases, hospitalizations, and deaths by age and geography over time in Michigan by integrating public health surveillance data, serial seroprevalence estimates, and genomic surveillance data. Burden estimates were used to examine how the risk of hospitalization and death varied over time by age and by SARS-CoV-2 variant.

## METHODS

### Cases

Confirmed cases of COVID-19 were taken from those reported to the Michigan Disease Surveillance System (MDSS) with cases reported in the Michigan Department of Corrections system excluded. MDSS data were accessed via a data use agreement between the University of Michigan and the Michigan Department of Health and Human Services. The IRB at the University of Michigan Medical School reviewed this project and determined it to be exempt secondary research for which consent is not required.

Age group- (0 to 17, 18 to 49, 50 to 64, and 65 plus years) and time-specific (March to May 2020, June to September 2020, October 2020 to February 2021, March to May 2021, and June to November 2021) adjustment factors were estimated in a model fit to case incidence data from MDSS and seroprevalence data from the state of Michigan from the CDC’s Nationwide Commercial Lab Seroprevalence study^8^. The model, adapted from Shioda et al., estimates cumulative incidence of infection from seroprevalence data while accounting for waning (Supplemental Figure 1)^6^. Age-group specific models were run to estimate the four time-specific adjustment factors using Markov Chain Monte Carlo sampling; parameter point estimates were taken as the median posterior sample, and 95% credible intervals (CrI) were taken as the 2.5th and 97.5th percentiles. As in Shioda et al., the time from illness onset to seroconversion was assumed to follow a Weibull distribution with mean 11.5 days and SD 5.7 days^9^. We also assumed that the average time from seroconversion to seroreversion (i.e. the duration a case would be detectable by serology) followed a Weibull distribution with mean 229.7 days (7.6 months) and SD 105.3 days. These parameters were estimated by fitting a Weibull distribution, using a weighted least squares method, to published data on the duration of seropositivity as measured by the Abbott ARCHITECT SARS-CoV-2 IgG immunoassay targeting the nucleocapsid protein (Supplemental Figure 2)^10,11^. This assay was used in the CDC’s Nationwide Commercial Lab Seroprevalence study to estimate seropositivity in Michigan^4^.

**Figure 1.**
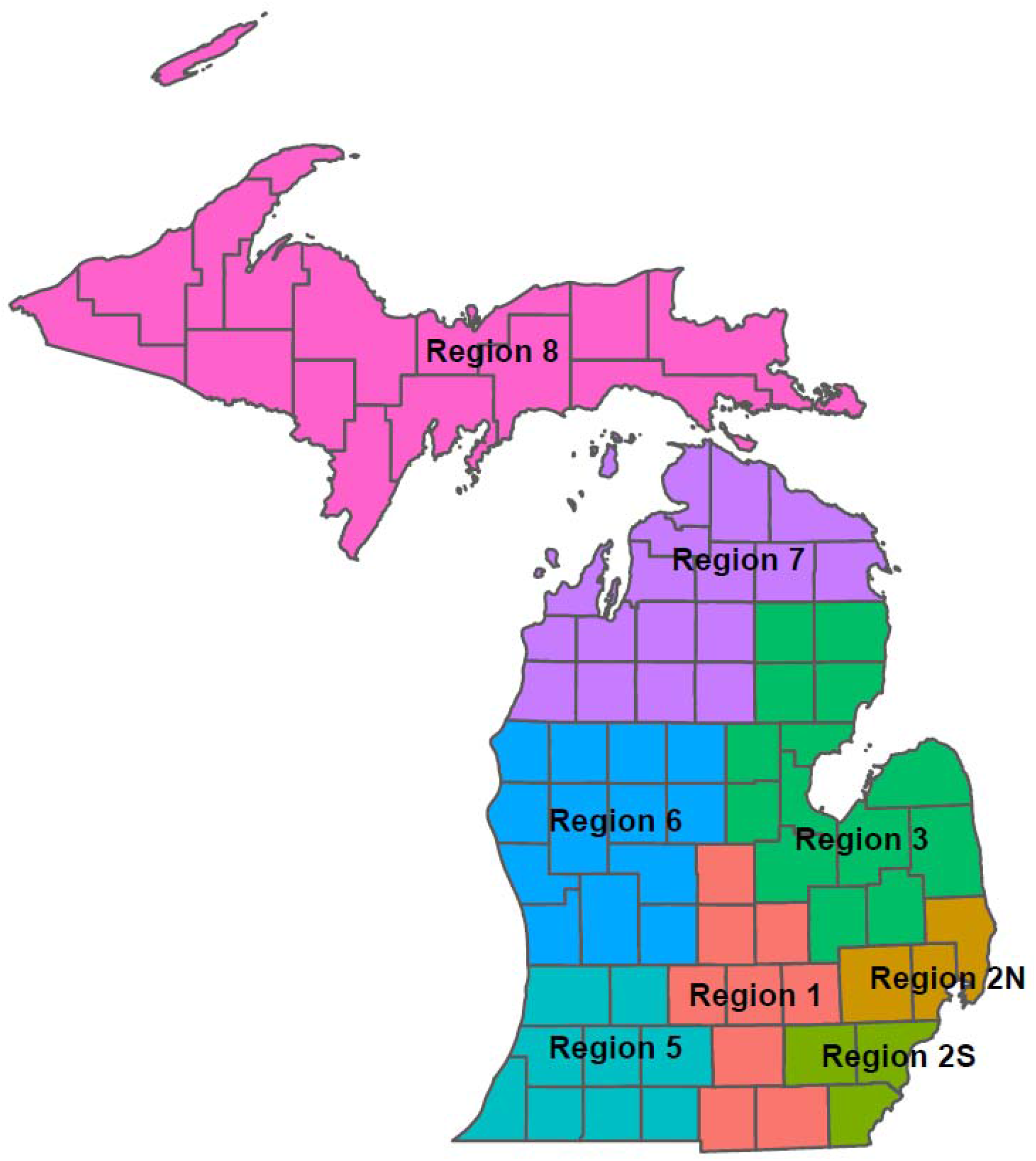
Map of Michigan Public Health Preparedness Regions.

**Figure 2.**
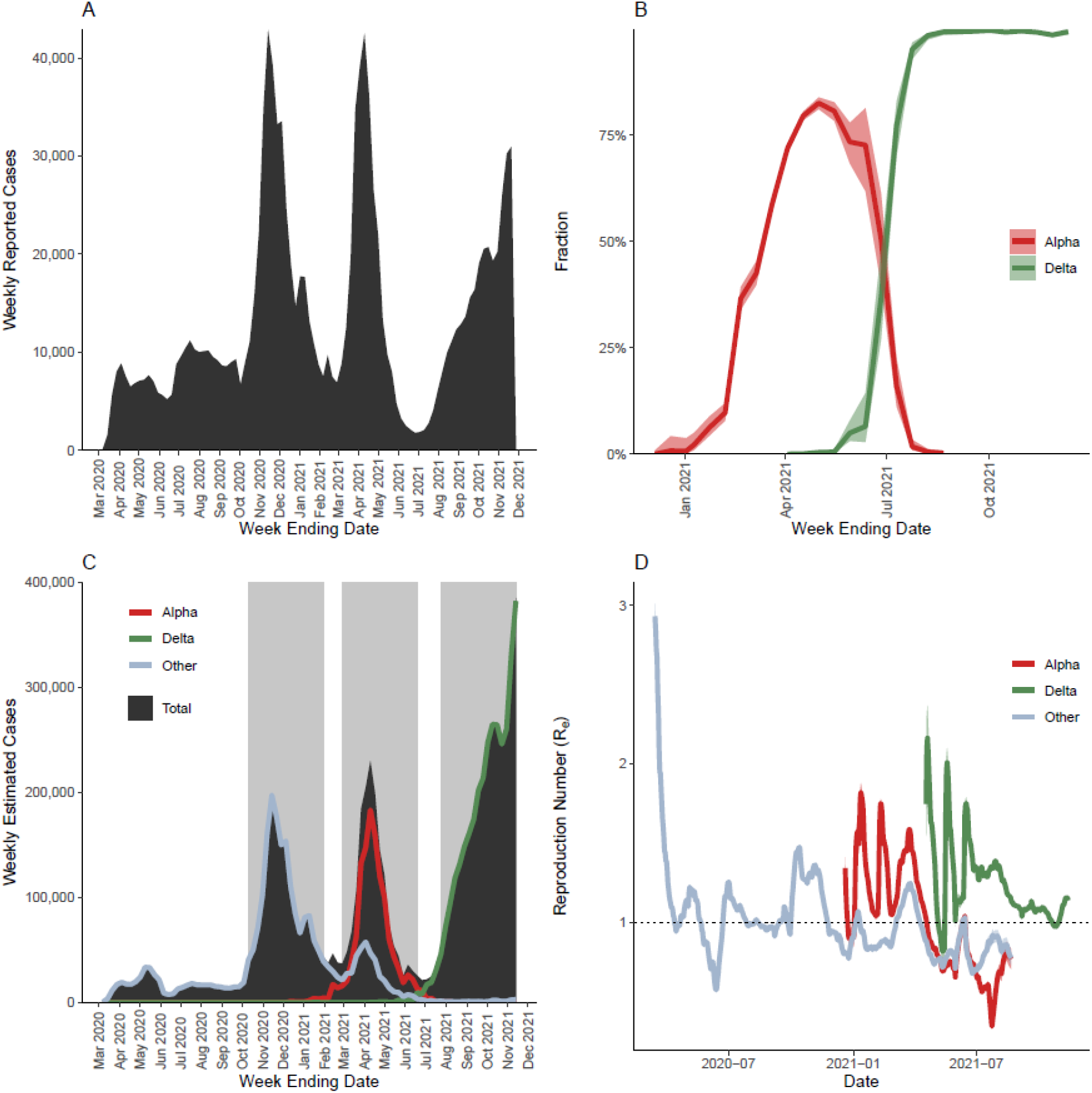
Epidemiology of the SARS-CoV-2 pandemic in Michigan. (A) Weekly reported confirmed and probable SARS-CoV-2 cases in Michigan. (B) Fraction of Alpha and Delta variant among characterized virus isolates. (C) Epidemic curve of estimated total, Alpha, Delta, and non-variant SARS-CoV-2 cases in Michigan; shaded gray areas highlight periods of analysis comparing Fall 2020 (Oct 11 - Jan 30), Spring 2021 (Feb 28 - Jun 19), and Fall 2021 (Jul 25 - Nov 13) waves. (D) Estimated reproduction number (R_e_) comparing Alpha, Delta, and non-variant viruses.

Daily cases from MDSS were then multiplied by the age- and time-specific adjustment factors and their 95% CrI to estimate a range of total infections. Adjusted total infections were then aggregated by age group and week to overall state, and public health preparedness region. Age groups used throughout this analysis are: 0 to 17, 18-19, 20-29, 30-39, 40-49, 50-59, 60-69, 70-79, and 80 plus years based on age group granularity in the available excess death data.

### Hospitalizations

Facility-level weekly adult and pediatric inpatient admissions for confirmed or suspected COVID-19 were identified from the HHS Protect COVID-19_Reported_Patient_Impact_and_Hospital_Capacity_by_Facility file^12^. Facilities were mapped to public health preparedness region (Figure 1), and overall weekly counts of admissions were aggregated to overall state and region levels. Hospitalized cases were also identified from MDSS data; because hospital admission status is ascertained at the time of case investigation and individuals may not have been admitted to the hospital yet, these data represent a severe underestimate of the total hospitalizations, but they do have information on age and public health preparedness region. For each week in each region, the number of hospitalized cases in each age group were divided by the total number of hospitalized cases in the state as a whole. The total number of weekly hospital admissions obtained from the HHS Protect data was then multiplied by the week- and region-specific age fraction obtained from MDSS to estimate the total number of hospitalizations in each age group, region, and week.

### Deaths

Weekly estimates of deaths were made using National Center for Health Statistics (NCHS) excess death estimates (Excess_Deaths_Associated_with_COVID-19 file^13^) and confirmed and probable COVID-19 deaths reported to MDSS with those reported in the Michigan Department of Corrections system excluded. For each week, the total count of deaths for the state was estimated as the total COVID-19 deaths reported to MDSS or the higher estimate of excess deaths from NCHS, whichever was higher. Using MDSS, the number of age group- and region-specific deaths in a week were divided by the total number of deaths in the state during that week. The total number of weekly deaths from the combined MDSS/NCHS data was then multiplied by the week- and region-specific age fraction obtained from MDSS to estimate the total number of deaths in each age group, region, and week.

### Variant Prevalence

Proportions of Alpha and Delta variant and ancestral lineage SARS-CoV-2 viruses among all characterized viruses in Michigan were obtained from covariants.org for 2-week periods^14^. Counts of cases, hospitalizations, and deaths were multiplied by the week-specific variant proportions to estimate the total number of cases, hospitalizations, and deaths attributable to Alpha, Delta, and ancestral viruses.

### Vaccination

The proportion of the population receiving at least 1 dose of vaccine by age and region was calculated by week from publicly available data reported to the Michigan Care Improvement Registry (Covid_Vaccine_Coverage_by_County_718469_7.xlsx^15^). Available vaccination data were available by the following age groups: 5 to 11, 12 to 15, 16-19, 20-29, 30-39, 40-49, 50-64, 65-74, 75 plus years. Vaccination counts were reassigned to the analysis age groups described under *Cases* as follows. If a vaccination age group spanned two analysis age groups, vaccination counts were assigned to each of the analysis age groups according to the proportion of age years contained in each age group. For example, 2/3 of the vaccinations in the 50 to 64 year vaccine age group were attributed to the 50 to 59 year analysis age group and 1/3 to the 60 to 69 year analysis age group. Vaccination counts from the 75 plus years vaccination age group were assigned to the 70 to 79 and 80 plus years analysis age groups according to the actual proportion of 75 plus year olds in Michigan who are also 80 or more years old (57%).

### Analysis

The 16 highest incidence weeks of the Fall 2020, Spring 2021, and Fall 2021 waves of COVID-19 in Michigan were compared. The Fall 2020 wave was defined from 10/11/2020 through 1/30/2021, the Spring 2021 wave was defined from 2/28/2021 through 6/19/2021, and the Fall 2021 wave was defined from 7/25/2021 through 11/13/2021. Delta variant transmission in Fall 2021 had not yet peaked at the time of analysis. Age-specific counts of estimated cases, hospitalizations, and deaths were plotted and compared across the waves, and by predicted variant status. Age- and region-specific proportions of cases who were hospitalized and proportions that died were also compared across waves. The effective reproduction number (Re) was estimated for Alpha, Delta, and ancestral lineage viruses over rolling 2-week intervals using the EpiEstim package with the Serial Interval specified as: mean_SI = 5.68, std_SI = 4.77^16,17^.

## RESULTS

From 10/11/2020 through 1/30/2021 359,061 confirmed cases were reported in Michigan, 285,528 confirmed cases were reported from 2/28/2021 through 6/19/2021, and 262,258 confirmed cases were reported from 7/25/2021 through 11/13/2021 (Figure 2A). After applying age and time specific adjustment factors (Supplemental Table 1), we estimated that there were 1,649,547 total cases in the first 16 weeks of the Fall 2020 wave, 1,594,954 total cases in the first 16 weeks of the Spring 2021 wave, and 3,329,748 total cases in the first 16 weeks of the Fall 2021 wave. Alpha variant viruses were first detected in December 2020 (Figure 2B). The proportion of sequenced viruses identified as Alpha variant rapidly increased to nearly 50% by late February / early March, and further increased to approximately 75% by early April. Delta variant viruses began to emerge in Michigan in April 2021 and accounted for nearly 100% of cases by mid-July 2021. Applying the estimated variant proportions to the estimated total weekly case time series, we estimated that there were 1,178,658 (74%) Alpha variant cases and 397,379 (25%) ancestral lineage cases in the Spring 2021 wave (Figure 2C). Ancestral lineage and Delta variant cases accounted for over 99% of infections in the Fall 2020 and Fall 2021 waves, respectively. The reproduction number for Alpha variant viruses was 12% (95% CrI: 12%,13%) higher than that of ancestral viruses, and the reproduction number for Delta variant viruses was 91% (95% CrI: 91%,92%) higher than that of Alpha variant viruses averaged over periods with overlapping circulation (Figure 2D).

Compared with the Fall 2020 wave, there was a higher burden of infection among the youngest Michigan age group, but lower burden among the over 55 year age groups in Spring 2021 (Figure 3). The relationship between burden and age appeared similar comparing Alpha and non-Alpha variant infections during Spring 2021; Alpha variant infections consistently accounted for approximately 2/3 of the total infections in each age group. The Fall 2021 Delta variant wave was estimated to have substantially higher burden in terms of infections than the 2 prior waves. Burden was high in all age groups, but young children and middle-aged adults were most heavily affected. In all waves, hospitalization and death was unlikely in the youngest age groups; individuals over 65 were most likely to be hospitalized and to die in all waves.

**Figure 3.**
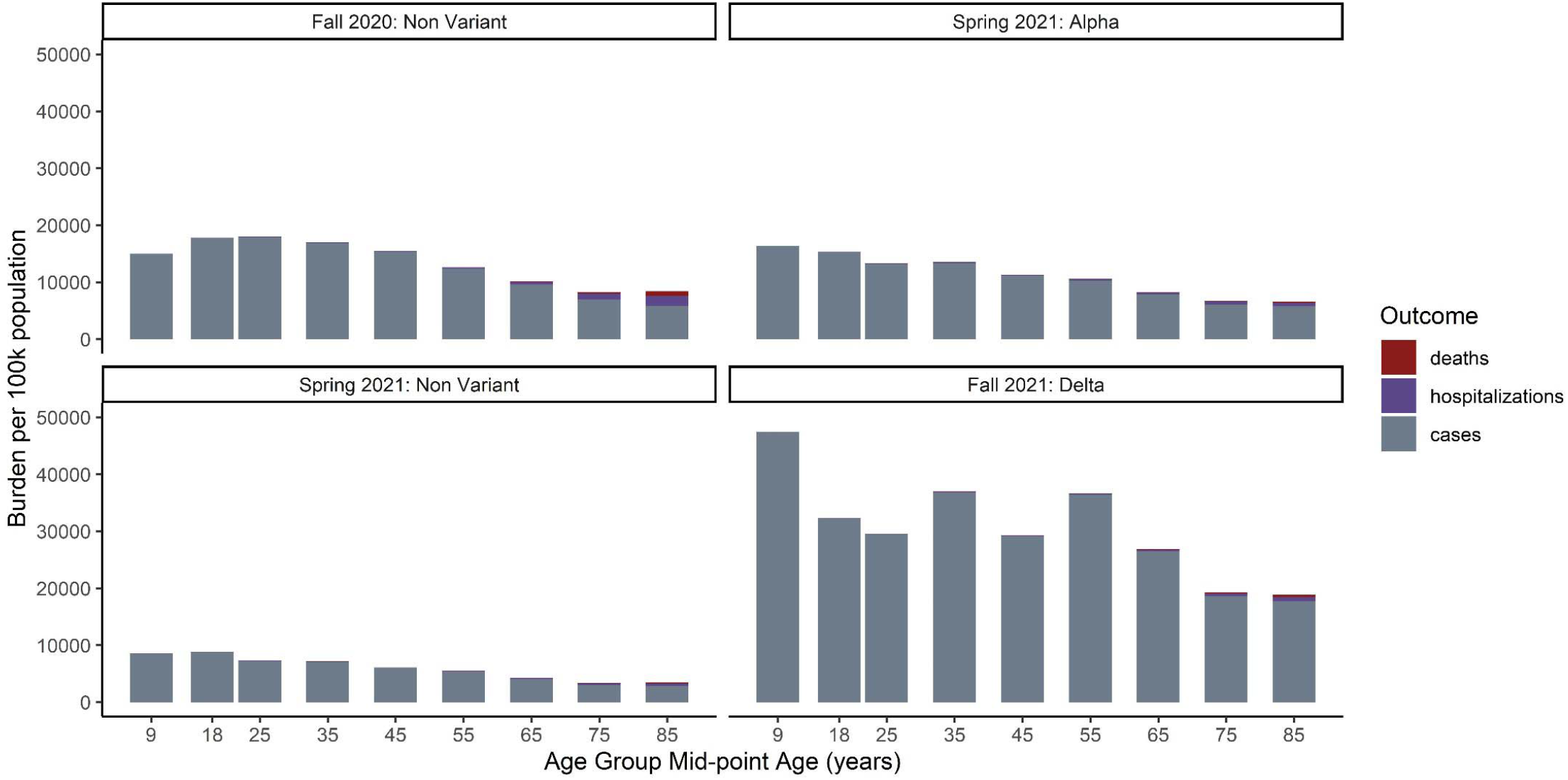
Estimated SARS-CoV-2 burden per 100,000 population by age in Michigan comparing estimated non-variant infections in Fall 2020 (Oct 11 - Jan 30), Alpha variant and non-variant infections in Spring 2021 (Feb 28 - Jun 19), and Delta variant infections in Fall 2021 (Jul 25 - Nov 13). The total height of the bars is the total number of cases in each age group i.e. hospitalizations and deaths were subtracted from the case counts. However, we do not know the proportion of hospitalized cases that died or the proportion of deaths that were not hospitalized so the combined height of those segments overestimates the total number of severe outcomes.

In all waves, the proportion of cases who were hospitalized and the proportion of cases who died in Michigan increased substantially with age. There was some suggestion that those in the middle age groups (20-29 through 50-59 years) were more likely overall to be hospitalized if infected in Spring 2021 than if infected in fall 2020 (Figure 4A). However, this did not appear to be Alpha variant specific; the likelihood of hospitalization did not vary by lineage in any age group in the spring (Figure 4B). Cases who were 80 years and older were more likely to be hospitalized (20% [95% CrI: 18%, 23%] vs 8% [95% CrI: 7%, 11%] vs 4% [95% CrI: 3%, 7%]) and to die (10% [95% CrI: 9%, 11%] vs 3% [95% CrI: 3%, 4%] vs 2% [95% CrI: 2%, 4%]) if infected in Fall 2020 than if infected in Spring 2021 or Fall 2021 (Figures 4A and 4C). This also did not appear to be driven by Alpha variant circulation in Spring 2021 as the proportion of cases who died was similar comparing Alpha and ancestral infections (Figure 3D). Across all age groups, those with Delta variant infection in Fall 2021 were less likely to be hospitalized or die than those with ancestral or Alpha variant infections in previous waves.

**Figure 4.**
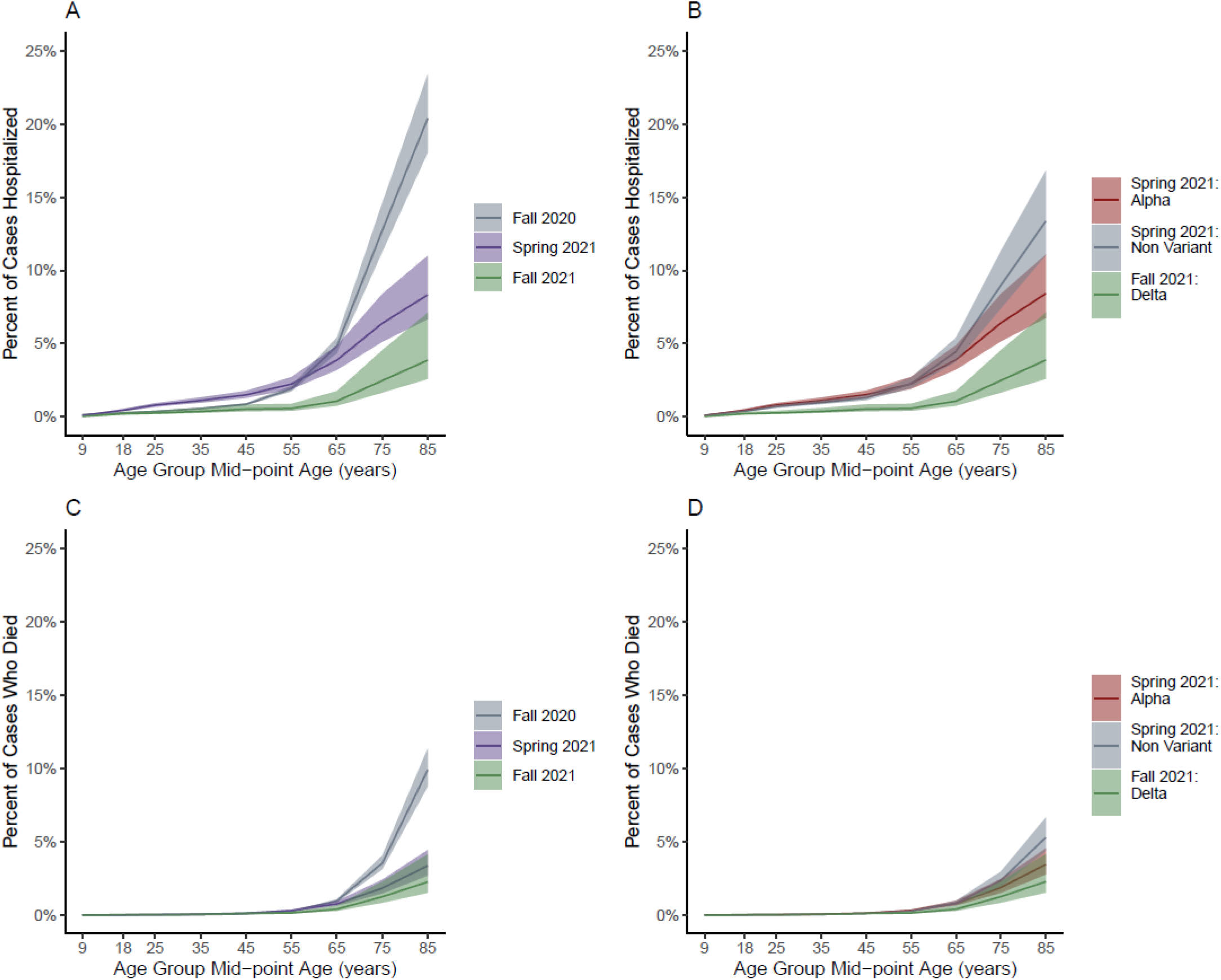
Percent of SARS-CoV-2 cases in Michigan who were hospitalized (A) comparing Fall 2020 (Oct 11 - Jan 30), Spring 2021 (Feb 28 - Jun 19), and Fall 2021 (Jul 25 - Nov 13) waves, and (B) comparing estimated Alpha variant and non-variant infections in Spring 2021, and Delta variant infections in Fall 2021. Percent of SARS-CoV-2 cases in Michigan who died (C) comparing Fall 2020, Spring 2021, and Fall 2021 waves, and (D) comparing estimated Alpha variant and non-variant infections in Spring 2021, and Delta variant infections in Fall 2021.

Cumulative incidence by age group was tightly clustered between 3% and 8% just prior to the Fall 2020 wave, with the 0 to 17 year old age group most likely (8%) and the 70-79 year old age group least like likely to have been previously infected (3%) (Figure 5A). Following the Fall 2020 wave, there was a wider range of cumulative incidence (13% to 28%) by age group with incidence generally decreasing with increasing age. The 18 to 19 year old age group was now most likely to have been previously infected (28%) and the 70 to 79 year old age group was least like likely to have been previously infected (13%) prior to the Spring 2021 wave, and we estimate that 22% of the total population had been infected. Following the Spring 2021 wave, overall cumulative incidence was 40% and ranged from 22% to 52% with similar patterns by age group. By November 12, 2021, overall cumulative incidence was 75% and ranged from 99% to 42% by age group. Cumulative incidence of infection did not vary as much by Michigan Public Health Preparedness Region as it did with age. However, the more rural Regions 7 and 8 consistently had the lowest cumulative incidence throughout the pandemic (Figure 5B).

**Figure 5.**
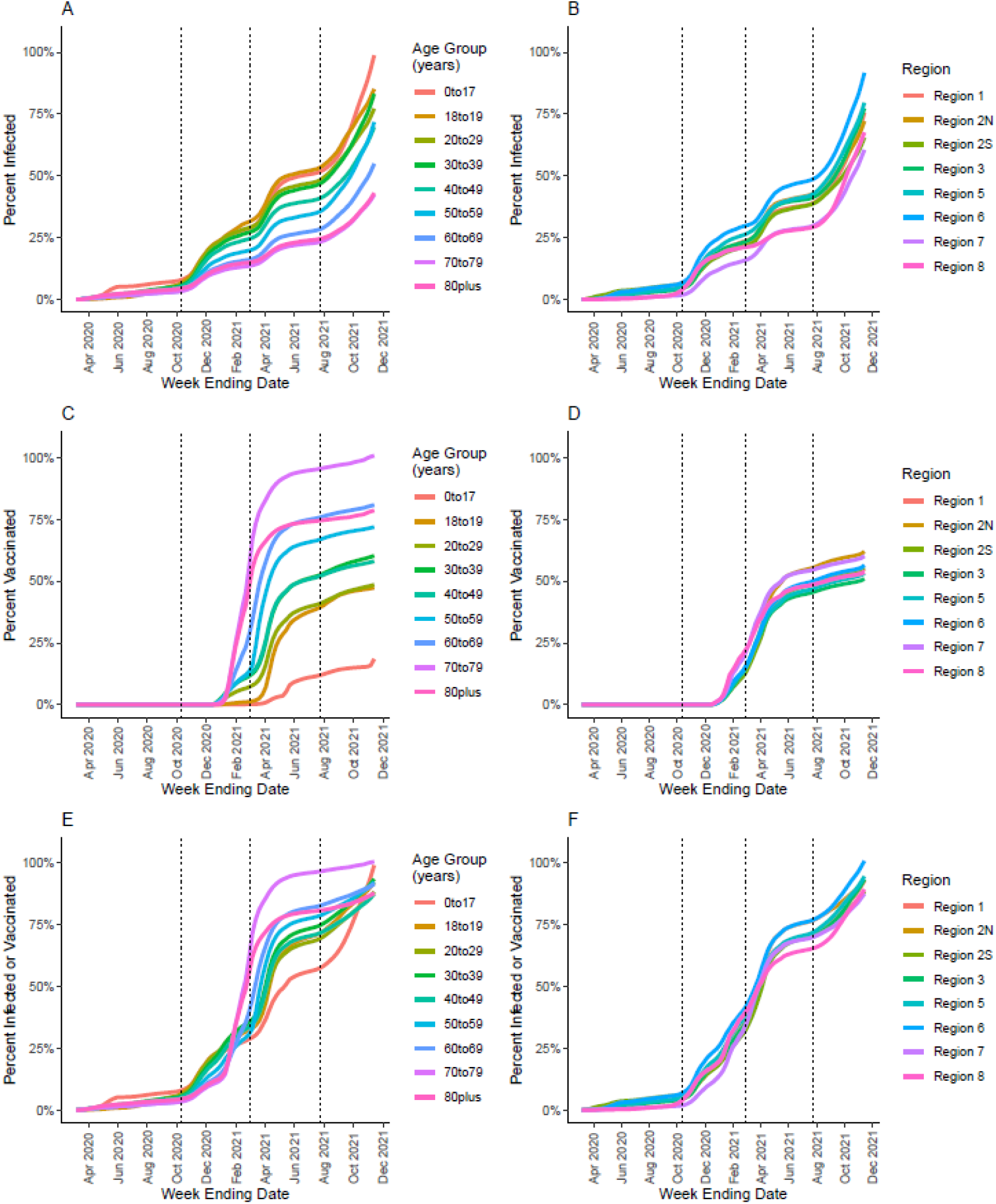
Cumulative percentage of Michigan population with SARS-CoV-2 infection, vaccination, or both by age group (A,C,E) and geographic region (B,D,F). Dotted vertical lines represent the start of the Fall 2020 (Oct 11 - Jan 30), Spring 2021 (Feb 28 - Jun 19), and Fall 2021 (Jul 25 - Nov 13) waves.

Substantial vaccine uptake occurred among the 60 to 69, 70 to 79, and 80+ year age groups in early 2021, with an estimated 47%, 55%, and 28% having at least one dose of vaccine, respectively, prior to the Spring 2021 wave (Figure 5C). Vaccination coverage increased in all age groups throughout Spring 2021 before slowing in the summer months, with final coverage ranging from >99% among 70 to 79 year olds to 19% among 0 to 17 year olds with coverage increasing with age. Vaccination coverage did not vary substantially by Public Health Preparedness Region, but uptake was initially faster in Regions 7 and 8 possibly reflecting an older population on average (Figure 4D). Proportions of individuals receiving at least one vaccination dose or previous infection by the end of Spring 2021 ranged from 96% among 70 to 79 year olds to 55% among 0 to 17 year olds and followed the same patterns as vaccination coverage (Figures 4E and 4F). By November 13, 2021, our model estimates that over 87% of Michigan residents in all groups had at least one vaccination dose and/or previous infection.

## DISCUSSION

We estimated substantial under-detection of SARS-CoV-2 infection that varied by age and time. Throughout the pandemic, infections in Michigan children were least likely to be detected. Under-detection among children was highest prior to May 2020 with an estimated 83 infections for every reported case. Detection improved to approximately 6 infections for every reported case in the summer of 2020 before worsening gradually over time to approximately 25 infections per report after June 2021. In contrast, infections among adults had much lower relative amounts of undetected infections throughout the pandemic. However, even with more accurate case counts in the adult age group, under-detection increased over time with the highest numbers of infections per reported case (8 to 14) after June 2021. Accounting for this under-detection, we estimated that the cumulative incidence of infection in Michigan reached 75% by mid-November 2021 with variability by age. Further accounting for vaccination, we estimate that the vast majority of Michiganders across all age groups had antigenic exposure to SARS-CoV-2 by mid-November 2021. These estimates can inform response and planning such as anticipating the scale of support services that will be needed for individuals with post-acute COVID-19 symptoms.

Following initial introductions of Alpha variant SARS-CoV-2, the state of Michigan experienced a large surge of infections in Spring 2021 when many other US states were experiencing declining incidence^7^. We estimate that Alpha variant infections accounted for approximately two thirds of the total infections during Michigan’s Spring 2021 wave. Consistent with the rapid replacement of non-Alpha variant viruses, we estimated that the effective reproduction number of the Alpha variant was 12% higher than ancestral viruses during the Spring. Alpha variant viruses were rapidly replaced by Delta variant viruses in the summer of 2021, and we estimated that the reproduction number of the Delta variant was almost twice as high as for Alpha in periods when their circulation overlapped.

These estimates are somewhat lower than previous reports that found the Alpha variant to be approximately 40% - 100% more transmissible than ancestral SARS-CoV-2 viruses^18,19^, and similar to reports of increased transmissibility of the Delta variant relative to Alpha^20,21^. Despite apparent reductions in the risk of severe outcomes given infection described previously, the continued emergence of more transmissible variants has stressed healthcare systems both through large patient volumes and infections among the healthcare workforce. This has been particularly highlighted by the ongoing wave of Omicron variant infections.

In the time since this analysis was carried out, infections with the Delta variant continued to rise in Michigan before being rapidly overtaken by the Omicron variant resulting in record high numbers of daily infection. Michigan’s experience in the winter of 2022 makes it clear that combined levels of prior infection and vaccination that exceed 80% are not sufficient to reach herd immunity. Suboptimal vaccine coverage, waning of natural and vaccine-induced immunity, and the emergence of more transmissible variants has facilitated ongoing transmission^22–24^. Indeed, it is unlikely that true herd immunity is sufficient to entirely end SARS-CoV-2 transmission in the near future, just as descendants of the 1968 and 2009 influenza pandemics continue to circulate today. This underscores the need for long term planning in policy, public health capacity, and research priorities as the pandemic continues. These results also suggest that non-pharmaceutical mitigation measures may be needed during times of high transmission going forward.

In general, we observed that the proportion of cases who were hospitalized and who died decreased from Fall 2020 to Spring 2021 to Fall 2021. Because of the ecologic nature of this analysis, we are unable to specifically attribute these general declines to variant-specific differences, vaccination, or some other cause. However, our results do not suggest any differences in the proportions of severe outcomes among the populations estimated to be infected with Alpha variant or ancestral viruses in the spring of 2021. Although differential severity has not been conclusively demonstrated, some studies have estimated Alpha and Delta are more severe than the ancestral SARS-CoV-2 strain, at least among unvaccinated individuals^25,26^. Estimates of SARS-CoV-2 vaccine effectiveness have been high against severe outcomes of infection^27–29^. This suggests that at least through the emergence of the Delta variant, age-specific reductions in severe outcomes were likely due to vaccination and improved treatment options over time.

A notable exception to the general trend of decreasing severity, is that adults less than 50 years of age appeared to be at increased risk of hospitalization in the spring of 2021 relative to the fall of 2020. While we were unable to examine individual level risk factors that might explain this difference, it is possible that patterns in vaccine uptake could confound differences in severity over time. Among those prioritized for early vaccination, younger adults with chronic conditions had similar low intention to vaccinate as “essential workers” generally^30^. Racial disparities in both vaccine uptake and severe outcomes of SARS-CoV-2 have also been demonstrated^31,32^. If healthier adults were more likely to be vaccinated, the remaining susceptible population may have been more likely to be hospitalized given infection. Factors associated with vaccine uptake warrant further investigation and consideration in analyses of differential severity.

We leveraged multiple sources of public health surveillance data and genomic surveillance to provide estimates of the overall burden, severity, and transmissibility of SARS-CoV-2 infection over time in Michigan. This was particularly challenging given the need to account for the changing vaccination and public health mitigation landscapes that had differential effects by age. The results should be interpreted in the context of multiple other limitations. 1) We were unable to account for reinfection, and prior infection was assumed to be independent of the likelihood of being vaccinated. 2) We relied on Michigan-specific data from the Nationwide Commercial Lab Seroprevalence study to calculate case adjustment factors to correct for under-detection. That seroprevalence study has its own limitations^4,8^, and the representativeness of its sample to the Michigan population as a whole is unclear. 3) The calculated case adjustment factors were sensitive to assumptions about the rate of antibody waning. We found that the average time from seroconversion to seroreversion could not be estimated simultaneously with the adjustment factors due to identifiability issues, so we specified the former parameter from the existing literature. It is possible that rates of antibody waning could differ by age which is something we were unable to account for. 4) The case adjustment factors were our main source of uncertainty in this analysis. There are other sources of uncertainty that we were unable to propagate or account for. 5) SARS-CoV-2 sequence data reported to GISAID may not reflect true community variant proportions, particularly shortly after emergence when sampling may be biased toward outbreaks.

Our results highlight the ongoing risk of periods of high SARS-CoV-2 incidence despite widespread prior infection and vaccination in the population. This underscores the need for long term planning for surveillance, vaccination, and other mitigation measures amidst continued response to the acute pandemic. The multiple streams of data on case incidence, infection outcomes, vaccine uptake, and genomic characterization that have facilitated the ongoing responses to the COVID-19 pandemic should be leveraged to facilitate ongoing surveillance and response and inform updates to SARS-CoV-2 vaccine composition and delivery schedule.

## Data Availability

All data produced in the present study are available upon reasonable request to the authors

## ACKNOWLEDGEMENTS

We thank Melissa Rolfes, Carrie Reed, Brendan Flannery, Miranda Delahoy, and Alicia Budd for extremely helpful conversations regarding methodology and preliminary results.

## SUPPLEMENT

**Supplemental Figure 1.**
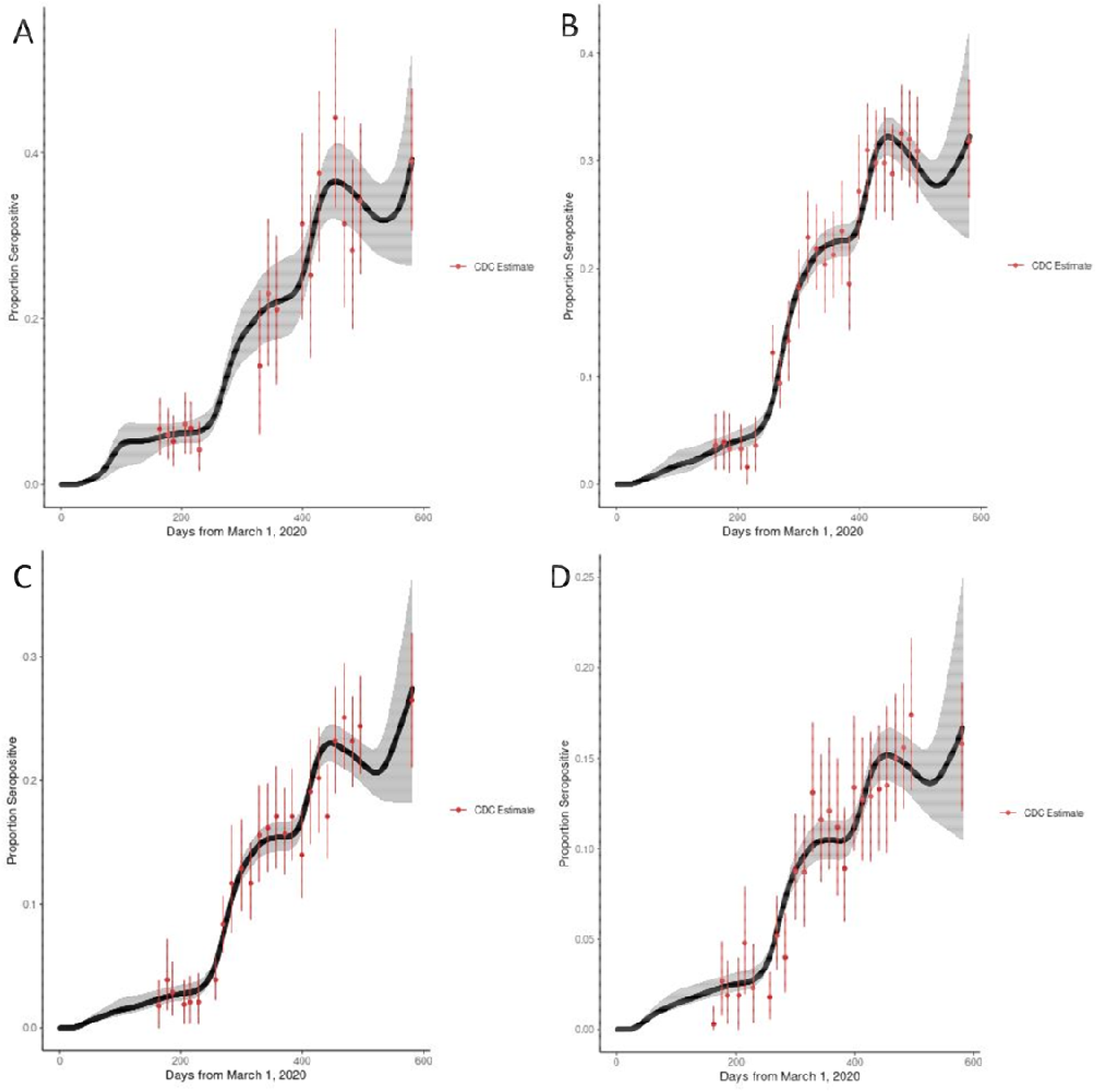
Estimated seropositivity over time among A) 0 to 17; B) 18 to 49; C) 50 to 64; and D) ≥65 year olds in Michigan (black line, 95% credible interval: gray area). Estimates and 95% confidence intervals from the CDC Nationwide Commercial Laboratory Seroprevalence Survey are in red.

**Supplemental Figure 2.**
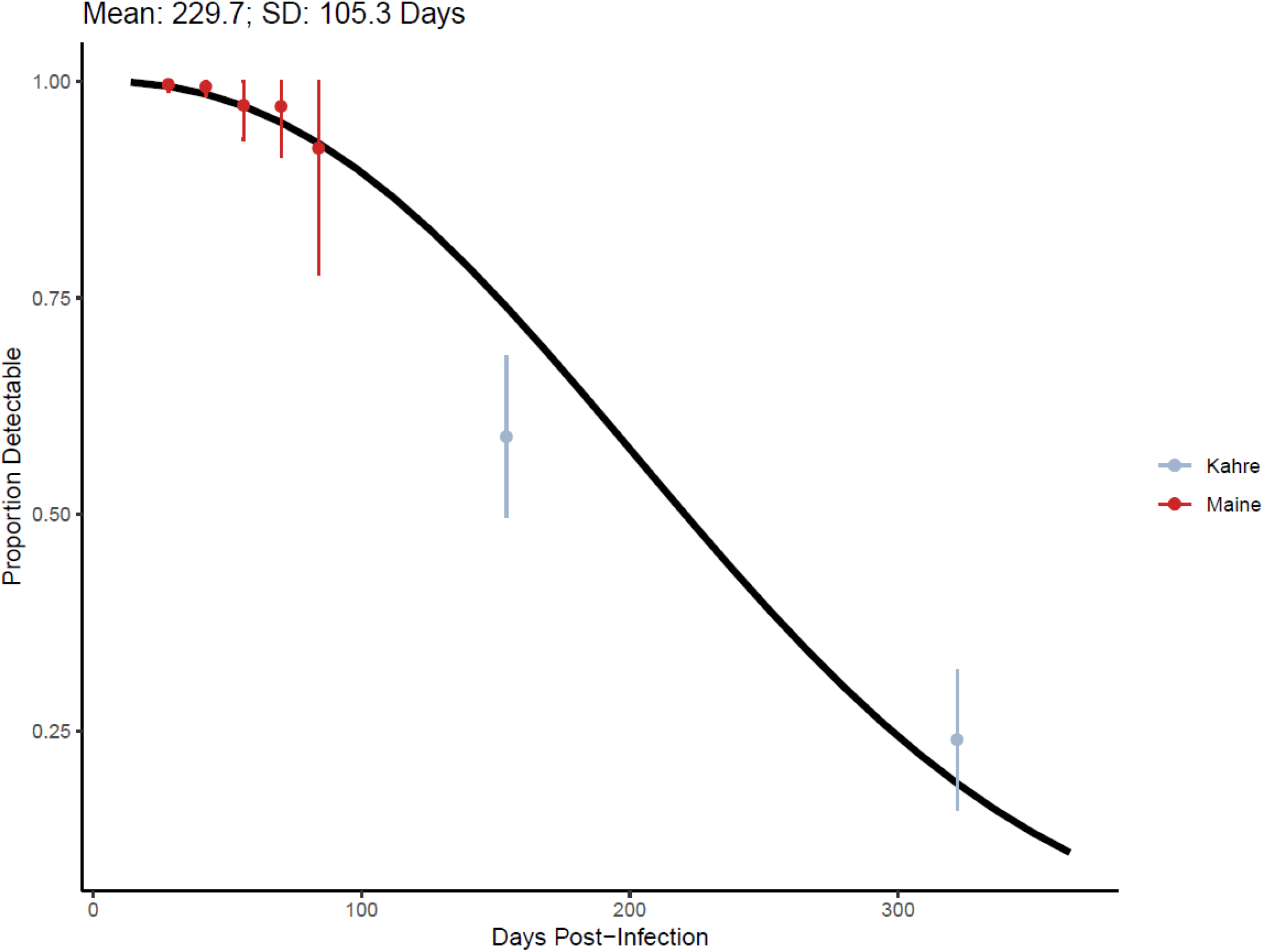
Probabability of detectable antibody to nucleocapsid protein by time following SARS-CoV-2 invection.

**Supplemental Table 1.** Case adjustment factors estimated from incidence and seroprevalence data.

|  | MAR - MAY 2020 | JUN - SEP 2020 | OCT 2020 - FEB 2021 | MAR - MAY 2021 | JUN - NOV 2020 |
| --- | --- | --- | --- | --- | --- |
| 0 to 17 years | 83.0 (35.3, 119.7) | 5.7 (1.3, 15.1) | 15.0 (11.3, 18.9) | 11.5 (7.8, 15.1) | 25.1 (12.6, 38.2) |
| 18 to 49 years | 2.1 (1.1, 3.7) | 1.4 (1.0, 2.1) | 4.3 (3.9, 4.6) | 4.3 (3.7, 4.9) | 9.8 (6.1, 13.7) |
| 50 to 64 years | 1.5 (1.0, 2.4) | 1.2 (1.0, 1.8) | 3.5 (3.2, 3.8) | 4.6 (3.8, 5.3) | 14.4 (9.3, 19.9) |
| 65+ years | 1.2 (1.0, 1.9) | 1.2 (1.0, 1.8) | 2.5 (2.2, 2.9) | 4.9 (3.8, 6.0) | 7.6 (4.2, 11.4) |

## References

1. Oran DP, Topol EJ. The Proportion of SARS-CoV-2 Infections That Are Asymptomatic. Ann Intern Med. 2021;174(5):655–662. doi:10.7326/M20-6976

2. Sah P, Fitzpatrick MC, Zimmer CF, et al. Asymptomatic SARS-CoV-2 infection: A systematic review and meta-analysis. PNAS. 2021;118(34). doi:10.1073/pnas.2109229118

3. Reese H, Iuliano AD, Patel NN, et al. Estimated Incidence of Coronavirus Disease 2019 (COVID-19) Illness and Hospitalization-United States, February-September 2020. Clin Infect Dis. 2021;72(12):e1010–e1017. doi:10.1093/cid/ciaa1780

4. Bajema KL, Wiegand RE, Cuffe K, et al. Estimated SARS-CoV-2 Seroprevalence in the US as of September 2020. JAMA Internal Medicine. 2021;181(4):450–460. doi:10.1001/jamainternmed.2020.7976

5. Havers FP, Reed C, Lim T, et al. Seroprevalence of Antibodies to SARS-CoV-2 in 10 Sites in the United States, March 23-May 12, 2020. JAMA Intern Med. Published online July 21, 2020. doi:10.1001/jamainternmed.2020.4130

6. Shioda K, Lau MSY, Kraay ANM, et al. Estimating the Cumulative Incidence of SARS-CoV-2 Infection and the Infection Fatality Ratio in Light of Waning Antibodies. Epidemiology. 2021;32(4):518–524. doi:10.1097/EDE.0000000000001361

7. Centers for Disease Control and Prevention. COVID Data Tracker. Centers for Disease Control and Prevention. Published March 28, 2020. Accessed February 14, 2022. https://covid.cdc.gov/covid-data-tracker/#trends_dailycases

8. Centers for Disease Control and Prevention. Nationwide COVID-19 Infection-Induced Antibody Seroprevalence (Commercial laboratories). Centers for Disease Control and Prevention. Published March 28, 2020. Accessed February 14, 2022. https://covid.cdc.gov/covid-data-tracker/#national-lab

9. Iyer AS, Jones FK, Nodoushani A, et al. Dynamics and significance of the antibody response to SARS-CoV-2 infection. medRxiv. Published online July 20, 2020:2020.07.18.20155374. doi:10.1101/2020.07.18.20155374

10. Maine GN, Lao KM, Krishnan SM, et al. Longitudinal characterization of the IgM and IgG humoral response in symptomatic COVID-19 patients using the Abbott Architect. J Clin Virol. 2020;133:104663. doi:10.1016/j.jcv.2020.104663

11. Kahre E, Galow L, Unrath M, et al. Kinetics and seroprevalence of SARS-CoV-2 antibodies: a comparison of 3 different assays. Sci Rep. 2021;11(1):14893. doi:10.1038/s41598-021-94453-5

12. US Department of Health & Human Services. COVID-19 Reported Patient Impact and Hospital Capacity by Facility. Accessed February 14, 2022. https://healthdata.gov/Hospital/COVID-19-Reported-Patient-Impact-and-Hospital-Capa/anag-cw7u

13. National Center for Health Statistics. Excess Deaths Associated with COVID-19. Published February 9, 2022. Accessed February 14, 2022. https://www.cdc.gov/nchs/nvss/vsrr/covid19/excess_deaths.htm

14. Hodcroft EB. CoVariants: SARS-CoV-2 Mutations and Variants of Interest. Published 2021. Accessed February 14, 2022. https://covariants.org/

15. Michigan Department of Health and Human Services. COVID-19 Vaccine Coverage by County. Published 2021. Accessed February 14, 2022. https://www.michigan.gov/documents/coronavirus/Covid_Vaccine_Coverage_by_County_718469_7.xlsx

16. Cori A, Ferguson NM, Fraser C, Cauchemez S. A New Framework and Software to Estimate Time-Varying Reproduction Numbers During Epidemics. American Journal of Epidemiology. 2013;178(9):1505–1512. doi:10.1093/aje/kwt133

17. Reed IG, Walker ES, Landguth EL. SARS-CoV-2 Serial Interval Variation, Montana, USA, March 1–July 31, 2020 -Volume 27, Number 5—May 2021 -Emerging Infectious Diseases journal - CDC. doi:10.3201/eid2705.204663

18. Davies NG, Abbott S, Barnard RC, et al. Estimated transmissibility and impact of SARS-CoV-2 lineage B.1.1.7 in England. Science. 2021;372(6538):eabg3055. doi:10.1126/science.abg3055

19. Volz E, Mishra S, Chand M, et al. Assessing transmissibility of SARS-CoV-2 lineage B.1.1.7 in England. Nature. 2021;593(7858):266–269. doi:10.1038/s41586-021-03470-x

20. Allen H, Vusirikala A, Flannagan J, et al. Household transmission of COVID-19 cases associated with SARS-CoV-2 delta variant (B.1.617.2): national case-control study. The Lancet Regional Health – Europe. 2022;12. doi:10.1016/j.lanepe.2021.100252

21. Scientific Pandemic Influenza Group on Modelling, Operational sub-group (SPI-M-O). SPI-M-O: Consensus Statement on COVID-19. Published June 2, 2021. Accessed February 14, 2021. https://assets.publishing.service.gov.uk/government/uploads/system/uploads/attachment_data/file/993321/S1267_SPI-M-O_Consensus_Statement.pdf

22. Sah P, Moghadas SM, Vilches TN, et al. Implications of suboptimal COVID-19 vaccination coverage in Florida and Texas. The Lancet Infectious Diseases. 2021;21(11):1493–1494. doi:10.1016/S1473-3099(21)00620-4

23. Levin EG, Lustig Y, Cohen C, et al. Waning Immune Humoral Response to BNT162b2 Covid-19 Vaccine over 6 Months. New England Journal of Medicine. 2021;385(24):e84. doi:10.1056/NEJMoa2114583

24. Wheatley AK, Juno JA, Wang JJ, et al. Evolution of immune responses to SARS-CoV-2 in mild-moderate COVID-19. Nat Commun. 2021;12(1):1162. doi:10.1038/s41467-021-21444-5

25. Twohig KA, Nyberg T, Zaidi A, et al. Hospital admission and emergency care attendance risk for SARS-CoV-2 delta (B.1.617.2) compared with alpha (B.1.1.7) variants of concern: a cohort study. The Lancet Infectious Diseases. 2022;22(1):35–42. doi:10.1016/S1473-3099(21)00475-8

26. Grint DJ, Wing K, Houlihan C, et al. Severity of SARS-CoV-2 alpha variant (B.1.1.7) in England. Clin Infect Dis. Published online September 6, 2021:ciab754. doi:10.1093/cid/ciab754

27. Thompson MG, Stenehjem E, Grannis S, et al. Effectiveness of Covid-19 Vaccines in Ambulatory and Inpatient Care Settings. N Engl J Med. 2021;385(15):1355–1371. doi:10.1056/NEJMoa2110362

28. Lewis NM, Naioti EA, Self WH, et al. Effectiveness of mRNA vaccines in preventing COVID-19 hospitalization by age and burden of chronic medical conditions among immunocompetent US adults, March-August 2021. J Infect Dis. Published online December 21, 2021:jiab619. doi:10.1093/infdis/jiab619

29. Nasreen S, Chung H, He S, et al. Effectiveness of COVID-19 vaccines against symptomatic SARS-CoV-2 infection and severe outcomes with variants of concern in Ontario. Nat Microbiol. Published online February 7, 2022. doi:10.1038/s41564-021-01053-0

30. Nguyen KH. COVID-19 Vaccination Intent, Perceptions, and Reasons for Not Vaccinating Among Groups Prioritized for Early Vaccination — United States, September and December 2020. MMWR Morb Mortal Wkly Rep. 2021;70. doi:10.15585/mmwr.mm7006e3

31. Mackey K, Ayers CK, Kondo KK, et al. Racial and Ethnic Disparities in COVID-19-Related Infections, Hospitalizations, and DeathsL: A Systematic Review. Ann Intern Med. 2021;174(3):362–373. doi:10.7326/M20-6306

32. Baack BN. COVID-19 Vaccination Coverage and Intent Among Adults Aged 18–39 Years — United States, March–May 2021. MMWR Morb Mortal Wkly Rep. 2021;70. doi:10.15585/mmwr.mm7025e2

